# Enhanced Recovery After Surgery reduced length of stay after colorectal surgery in a small rural hospital in Ontario

**DOI:** 10.1101/2022.03.23.22272850

**Authors:** Hector A. Roldan, Andrew R. Brown, Jane Radey, John C. Hogenbirk, Lisa R. Allen

**Affiliations:** Chief of Surgery, Muskoka Algonquin Healthcare, 100 Frank Miller Drive, Huntsville, Ontario, P1H 1H7, Associate Professor, Northern Ontario Medical School, 935 Ramsey Lake Road, Sudbury, ON, Canada P3E 2C6; Anesthesiologist, Muskoka Algonquin Healthcare, 100 Frank Miller Drive, Huntsville, Ontario, P1H 1H7, Assistant Professor, Northern Ontario Medical School, 935 Ramsey Lake Road, Sudbury, ON, Canada P3E 2C6; Register Nurse, RNFA, CPN(C), Muskoka Algonquin Healthcare, 100 Frank Miller Drive, Huntsville, Ontario, P1H 1H7; Research Tutor, Postgraduate Medical Education Office, Northern Ontario School of Medicine, Associate Director, Centre for Rural and Northern Health Research, Laurentian University, 935 Ramsey Lake Road, Sudbury, ON, Canada P3E 2C6; Research Coordinator, Huntsville Physicians, South Muskoka and Parry Sound Local Education Groups, Muskoka Algonquin Healthcare, 100 Frank Miller Drive, Huntsville, Ontario, P1H 1H7

**Keywords:** Enhanced Recovery After Surgery (ERAS), length of stay (LOS), hospitals, rural, colorectal surgery, Ontario, perioperative care

## Abstract

**Background:** Enhanced Recovery After Surgery (ERAS) programs include preoperative, intraoperative and postoperative clinical pathways to improve quality of patient care while reducing length of stay and readmission. This study assessed the feasibility and outcomes of an ERAS protocol for colorectal surgery implemented over two-years in a small, resource-challenged rural hospital.

**Study design:** A prospective cohort study used retrospectively matched controls to assess the effect of ERAS on LOS in patients undergoing colorectal surgery in a small rural hospital in northern Ontario, Canada. ERAS patients were matched to two patients in the control group based on diagnosis, age and gender. Patients had open or laparoscopic colorectal surgeries, with those in the intervention group treated per ERAS protocol and given instructions on pre- and post-operative self-care.

**Results:** Most ERAS patients reported adherence to ERAS protocols prior to surgery. Approximately one quarter of patients chose not to complete the postoperative survey. Of those who completed the survey, adherence to protocol was strongest for chewing gum in the days after surgery. Most patients were sitting in a chair for their afternoon meal by the first day and most were walking down the hallway by the second day. The control and ERAS patient groups did not differ significantly (p≥0.07) in age (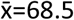years, sd=13.1), gender (52% male), nor in the Canadian Classification of Health Interventions 5-character code. The control group significantly higher (p<0.001) malignant neoplasm of colon (C18, 69% vs 35%), and significantly lower malignant neoplasm of rectum (C20, 0% vs 5%), relative to the ERAS group. The control group had an average ln-transformed LOS that was significantly longer (exponentiated as 1.7 days) than ERAS patients (t-test, p<0.001).

**Conclusion:** This study found that ERAS could be implemented in a small rural hospital and provided evidence for a reduced LOS of approximately two days.

## Background

Enhanced Recovery After Surgery (ERAS) programs consist of preoperative, intraoperative and postoperative clinical pathways to improve the quality of patient care while reducing the length of stay, readmission rates and reduce the economic impact on the institution (1–6). By following the 15 – 20 interventions defined by ERAS, many large centers have shown significant improvement in patient outcomes, fewer surgical site infections and lower rates of hospital acquired infection (2–4). However, evidence is sparse for the effectiveness of ERAS in smaller, rural hospitals (7). This study reports on the feasibility and selected outcomes of implementing the ERAS program in a small rural hospital located in an underserved region of Ontario, Canada.

ERAS programs use evidence-based medicine to challenge traditional surgical practices; strict fasting protocols replaced by carbohydrate loading, control and optimal goal-directed fluid therapy during surgery, advances in anesthesia allow catered approaches to minimize opioid use and early mobilization after surgery is encouraged (6–9).

ERAS also highlights the need for patient engagement in their own healing. Patients appreciated playing a role in their recovery and were highly satisfied with all aspects of their procedure such as physician skill level (technical and interpersonal), preoperative patient education and availability of staff to the patients (6,10,11).

Patient education is a key component from the preoperative stage through to post-operative follow-up (6,12). Patients are encouraged to take some responsibility for their post-surgical outcomes (13), particularly related to smoking cessation; smoking increases risk factors for wound healing, anastomotic leak, perioperative stroke and myocardial infarction. Consistent and correct information is crucial as demonstrated by previous research where 90% of older adults adhere more strongly to ERAS protocols when time is taken to ensure the patients understand the guidelines (10,11,13).

Conventionally, patients preparing to undergo gastrointestinal surgery would be in a fasting state for a minimum of eight hours to reduce the risk of aspiration pneumonia (14,15). Additionally, patients would undergo a bowel preparation, which may increase the risk of dehydration, particularly in the elderly (6,14). Patients that smoke, have functional dyspepsia, psychological stress or have an increase in female hormones are at increased risk for delayed gastric emptying (14).

ERAS preoperative procedures focus on patient engagement and optimal preparation for their surgical procedure in four key areas; breathing (smoking cessation), movement (exercise), nutrition and expectations (clear surgery date) (8,12). Intraoperatively the patient is maintained at the ideal anesthesia depth, has active warming, and goal-directed fluid therapy, particularly for high-risk patients (11). Patients are risk-stratified for nausea and vomiting and are given pre-emptive medication accordingly. Postoperatively, pain is managed with multi-modal therapy, minimizing opioid use ; narcotic use is a rate-limiting step in patients regaining bowel function, which directly influences LOS and can result in further complications (5,8,9). Epidural anesthesia is often part of this approach. Nasal gastric tubes, bladder catheters, drains and intravenous fluid are used sparingly and removed as soon as possible (8). Enteral feeding and early mobility are introduced as soon as feasible after surgery and routine screening for delirium is conducted for older adults (11).

Studies overwhelmingly suggest that adherence to the entire pathway produces the best patient outcomes (1,7,8) and highlights the need for healthcare professionals to work as a multidisciplinary team (2,7,11). The program requires input and support from all layers within the facility: hospital administrators and senior leadership, clinicians including surgeons, anesthesiologists and nurses, and allied health professionals such as physiotherapists and dietitians (3,16,17).

Successful execution of ERAS requires substantial changes from the traditional methodologies for gastrointestinal surgeries. While ERAS protocols have been in place in urban centers for several years, this may be a challenge in rural hospitals, which have fewer resources (7). The goal of this project was to determine the feasibility of implementing an ERAS protocol for gastrointestinal procedures over two years in a small rural hospital and to evaluate its impact on patient outcomes, LOS, morbidity and readmission rate.

## Methods

### Study design and setting

A prospective cohort study, using retrospectively matched controls, was used to assess the effect of ERAS on LOS in patients undergoing colorectal surgery in a small rural community hospital situated in northern Ontario. Huntsville, Ontario has a stable population of 6482 and a catchment area of just under 20,000 permanent residents quadrupling seasonally with tourists. Seniors represent 27% of the population as it is also a retirement destination (Statistics Canada, 2016). The local hospital, HDMH (one site of MAHC), has 37 acute care beds, dedicated for adult care. Children requiring hospitalization are referred elsewhere for pediatric inpatient services. This study received ethics approval from the Laurentian University Research Ethics Board (file number 2015-02-02) on April 10, 2015.

The surgical team involved the primary investigator HR, and two other surgeons, JM and RK. The study educator and surgical assistant is a Registered Nurse First Assistant (RNFA). Anesthesia for all surgeries was overseen by AB.

All patients undergoing routine colorectal surgery, either benign or malignant disease, were eligible for the ERAS project. Consent for patients undergoing colorectal surgery was attained as per normal procedure in the surgeon’s office. Patients were educated regarding the surgical procedure and expectations after which they had the opportunity to ask questions and have any aspect clarified. Family members were included when possible. Smoking cessation was mandatory four weeks prior to all ERAS procedures with patients receiving support aids if necessary. It was clarified that this was vital in patient recovery for ERAS procedures and surgery would be rescheduled if patients were unable to cease smoking. Participation was offered to all eligible patients between November 1, 2015 to November 1, 2017. All patients who were invited to participate, enrolled in the study. Consent was obtained for 47 patients.

### ERAS Protocol

The RNFA and, when required, the on-call anesthetist conducted patient education sessions in the day surgery unit. These visits lasted one to two hours. ERAS patients received detailed information about how to prepare for surgery, what they would need in the hospital and at home for their post-surgical care. Patients were sent home with the ERAS Patient Education Booklet.

Traditionally, patients would only attend the hospital pre-surgically if a consult was required. Surgical instructions and pre-surgical medications would be provided by their surgeon when the procedure was booked.

ERAS patients were advised to consume two carbohydrate drinks prior to their surgery (the night before and four hours before surgery). They were asked to chew gum as soon after recovery as possible and were encouraged to start eating solid food and drinking immediately after surgery. Mobility was promoted the night immediately following surgery by having patients sit and dangle their legs over their bed. Short walks were encouraged the day after surgery.

Traditionally patients are advised to take nothing by mouth from the evening prior to surgery until the day after surgery, though clear fluids are permitted after patients leave recovery. Patients often remain in their bed for an extended period, until they feel well enough to walk.

When ERAS patients were seen by their surgeon two to four-weeks prior to their surgery, they were instructed to optimize their nutrition and improve their cardiovascular activity. Smoking cessation was required four weeks prior to surgery. Prescriptions were provided for oral antibiotics and bowel preparation at the surgeon’s discretion. Upon completion of their presurgical appointment, patients could be referred for a preoperative anesthesia and/or internal medicine consultation if not already done. Patients were asked to bring their ERAS patient handbook to the hospital with them on the day of their surgery with the preoperative questionnaire completed in advance.

*On the day of surgery*, patients were instructed to fast after midnight, except for clear fluids as desired and a mandated liquid carbohydrate load four hours prior to surgery. Once in the operating room, a surgical checklist was completed as per routine hospital procedure. Intravenous antibiotics, to help prevent surgical site infection, were initiated one hour prior to surgery, and deep-vein thrombosis prophylaxis treatment, including compression stockings and sequential compression devices, was used. Patients were warmed during surgery with an air blanket device to maintain their body temperature while actively monitoring their temperature throughout.

Each ERAS patient received thoracic epidural anesthesia prior to anesthetic induction. Induction of general anesthesia was done via usual technique with opioids, propofol and rocuronium dosed individually by the attendant physician. Immediately after induction, an esophageal doppler probe, which generates individualized, estimated real-time cardiac output, was placed to facilitate intraoperative goal-directed fluid therapy. Patients were monitored in the usual fashion during surgery and transferred to the ICU for monitoring and care after surgery.

*After surgery*, while still in hospital, patients tracked their progress in their patient handbook. Many wrote additional notes and comments in the margins of their handbook about their experience, interaction with staff or how they were feeling. The patient handbook was left with the nursing staff to be collected by the research team when the patient was discharged. ERAS post-operative recommendations included early mobilization after surgery, chewing gum daily, early return to normal diet and the optimal use of pain management.

### Data collection and analyses

Patients were asked to complete the ERAS patient handbook pre-and postoperatively. Questions were asked about the patient’s role and expectations for recovery. Patients were also asked to report their perceived pain using a 10-point visual analogue scale with 0 being no pain and 10 being the highest pain they had experienced. Data were also collected from the hospital’s EMR including: sex, age (years), most responsible diagnosis (coded by International Statistical Classification of Diseases and Related Health Problems, 10^th^ Revision, Canada, ICD-10-CA), principal surgical procedure (coded by the Canadian Classification of Health Interventions, CCI), and LOS, in days. Additional data were collected for patients who were enrolled in the ERAS program from November 2015 to November 2017. These data included presence of ileus, vomiting/nausea, urinary retention, wound infection or dehiscence, deep vein thrombosis, pneumonia, anastomotic leak and readmission. Of the 47 patients recruited to the study, seven patients were removed at the discretion of the attending surgeon due to significant post-operative complications including complicated ileus, substantial nausea and vomiting and a case of abdominal dehiscence that required re-suturing. These complications were outlined in the ERAS order set at the beginning of the study.

An initial control group of 91 patients was obtained from the EMR for two years prior to implementation of the ERAS program. Patients were matched on diagnosis, age and sex. A total of 11 patients were removed from the control group because they experienced similar complications to those patients that were excluded from the study (significant ileus, vomiting and diarrhea, surgical site complications).

The primary outcome was LOS, measured in whole days, with 80 patients in the control group and 40 patients in the ERAS group. The choice of statistical procedures was informed by Chazard et al. (2017) who recommended Student’s t-test on logarithmic transformed data or the Mann-Whitney (Wilcoxon) test for two independent groups. Chazard’s recommendations, developed for equal sample size, were assumed to apply to this study with twice as many patients in the control group than in the ERAS group. We also used Student’s t-test and Fisher’s test (using exact methods or Monte Carlo methods based on 10,000 randomly sampled tables) to look for differences in patients’ age and sex, as well as ICD-10-CA and CCI codes between the control and ERAS group. We used McNemar tests to look for differences in self-reported pain scores from the night following the surgery to days 1, 2 and 3 post-surgery. All analyses were conducted with IBM SPSS Statistics for Windows, Version 24.0 (Armonk, NY: IBM Corp.).

## Results

### ERAS patients, protocol, and outcomes

Exactly 55% of ERAS patients were male and were somewhat older, though not statistically significant, than female patients in this group (Chi-squared test, p=0.57) (**Table 1**). At least 57% of patients had adhered to preoperative instructions to drink bowel preparation and bring chewing gum with them (**Table 2**). Patients’ recall of what was expected of them and their expected LOS was 65% or higher with one exception; only 48% (19 of 40) of patients recalled being informed that they would be able to consume solid foods the day after surgery.

**Table 1.** Age-sex distribution of 40 ERAS patients

|  | Male |  | Female |  | All Patients |  |
| --- | --- | --- | --- | --- | --- | --- |
| Age (years) | Count | Percent | Count | Percent | Count | Percent |
| <60 | 6 | 27% | 8 | 44% | **14** | **35%** |
| 60-69 | 4 | 18% | 4 | 22% | **8** | **20%** |
| 70-79 | 7 | 32% | 4 | 22% | **11** | **28%** |
| 80-89 | 5 | 23% | 2 | 11% | **7** | **18%** |
| **Sub-total** | **22** | **100%** | **18** | **100%** | **40** | **100%** |

**Table 2:** ERAS Procedural Compliance as reported by patients pre-and post-operatively

| Question | Yes | No | No Answer |
| --- | --- | --- | --- |
| Did you drink bowel preparation? | 28<br>70% | 3<br>8% | 9<br>23% |
| Did you bring chewing gum with you? | 24<br>57% | 1<br>2% | 17<br>40% |
| Were you informed that you are expected to dangle your legs out of bed within four hours of surgery? | 26<br>65% | 0<br>0% | 14<br>35% |
| Were you informed that you are expected to chew gum after surgery to help you pass gas? | 26<br>65% | 0<br>0% | 14<br>35% |
| Were you informed that you are expected to eat your meals in a chair, out of bed? | 26<br>65% | 0<br>0% | 14<br>35% |
| Were you informed that you are able to consume solid foods the day after surgery? | 19<br>48% | 5<br>13% | 16<br>40% |
| Were you informed that your LOS is expected to be three days (colon) or four days (rectal)? | 30<br>75% | 0<br>0% | 10<br>25% |
| Were you encouraged to drink a high carbohydrate drink the night before your surgery? | 31<br>78% | 0<br>0% | 9<br>23% |
| Were you encouraged to drink a high carbohydrate drink two hours before your surgery? | 31<br>78% | 0<br>0% | 9<br>23% |
| **Night of surgery (with help)** |  |  |  |
| I sat at the side of my bed for 10 – 15 minutes (with help). | 18<br>45% | 9<br>23% | 13<br>33% |
| I did deep breathing exercises 10 times per hour when I was awake. | 22<br>55% | 10<br>25% | 8<br>20% |
| I was offered sips of clear fluids | 29<br>73% | 3<br>8% | 8<br>20% |

| Days After Surgery | Day 1 |  |  | Day 2 |  |  | Day 3 |  |  |
| --- | --- | --- | --- | --- | --- | --- | --- | --- | --- |
| Question | Yes | No | No Answer | Yes | No | No Answer | Yes | No | No Answer |
| I sat in the chair for my morning meal. | 15<br>38% | 13<br>33% | 12<br>30% | 23<br>58% | 6<br>15% | 11<br>28% | 23<br>58% | 7<br>18% | 10<br>25% |
| I sat in the chair for my afternoon meal. | 21<br>53% | 9<br>23% | 10<br>25% | 22<br>55% | 6<br>15% | 12<br>30% | 22<br>56% | 5<br>13% | 12<br>30% |
| I sat in my chair at other times throughout the day. | 16<br>40% | 9<br>23% | 15<br>38% | 23<br>58% | 4<br>10% | 13<br>33% | 15<br>38% | 4<br>10% | 21<br>53% |
| I walked down the hall at least once. | 17<br>41% | 9<br>22% | 15<br>37% | 23<br>58% | 2<br>5% | 15<br>38% | 23<br>58% | 2<br>5% | 15<br>37% |
| I had nothing to eat or drink | 0<br>0% | 30<br>75% | 10<br>25% | 3<br>8% | 6<br>15% | 31<br>78% | 2<br>5% | 6<br>15% | 32<br>80% |
| I had liquids to eat/drink. | 29 | 0 | 11 | 26 | 1 | 13 | 23 | 0 | 17 |
| Question | Yes | No | No Answer | Yes | No | No Answer | Yes | No | No Answer |
|  | 73% | 0% | 28% | 65% | 3% | 33% | 58% | 0% | 43% |
| I had solid food. | 17<br>43% | 4<br>10% | 19<br>48% | 18<br>45% | 3<br>8% | 19<br>48% | 19<br>38% | 3<br>8% | 28<br>70% |
| I chewed gum in the morning. | 28<br>72% | 2<br>5% | 9<br>23% | 28<br>72% | 3<br>8% | 8<br>21% | 25<br>63% | 3<br>8% | 12<br>31% |
| I chewed gum in the afternoon. | 31<br>78% | 1<br>3% | 8<br>20% | 29<br>73% | 2<br>5% | 9<br>23% | 22<br>55% | 3<br>8% | 15<br>38% |
| I chewed gum in the evening. | 27<br>68% | 2<br>5% | 11<br>28% | 29<br>73% | 3<br>8% | 8<br>20% | 19<br>48% | 3<br>8% | 18<br>45% |
| My catheter came out today. | 13<br>33% | 16<br>40% | 11<br>28% | 14<br>35% | 11<br>28% | 15<br>38% | 11<br>28% | 11<br>28% | 18<br>45% |
| I am peeing on my own. | 9<br>23% | 20<br>50% | 11<br>28% | 19<br>48% | 10<br>25% | 11<br>28% | 22<br>55% | 4<br>10% | 14<br>35% |
| I am passing gas. | 16<br>40% | 14<br>35% | 10<br>25% | 21<br>53% | 9<br>23% | 10<br>25% | 24<br>60% | 3<br>8% | 13<br>33% |

On the night following surgery, 45% of the patients dangled their legs from the bed with help, 55% completed their breathing exercises and 73% were offered clear fluids (**Table 2**). The day after surgery between 38% and 53% of patients consumed breakfast or lunch while sitting in their chair, 73% reported consuming liquids and 78% chewed gum at least once. Only 23% indicated they were ‘peeing on their own’, while 40% reported they were passing gas. On day two post-surgery, 56% of patients ate one meal in their chair and reported walking down the hall at least once, 45% were consuming solid food, 73% were chewing gum, 47% were ‘peeing on their own’ and 47% were passing gas. Day three findings were like day two.

The night of the surgery, 25% of patients reported moderately high pain (6–7) and 12% reported high pain (8–10) (**Table 3**). On the day following surgery, 20% of patients reported moderately high pain and 25% reported high pain. On days two and three after surgery, patients indicated a trend towards lower pain, though these day-to-day trends were not statistically significant (McNemar test, p>0.27).

**Table 3.** Number of ERAS patients and their self-reported daily pain measurement score. *†

| Time relative to Surgery | Pain level 0 | Pain level 1-3 | Pain Level 4-5 | Pain level 6-7 | Pain level 8-10 | No Answer |
| --- | --- | --- | --- | --- | --- | --- |
| First Night after surgery | 5<br>12% | 4<br>10% | 9<br>22% | 10<br>24% | 5<br>12% | 8<br>20% |
| Day 1 after surgery | 2<br>5% | 3<br>8% | 6<br>15% | 8<br>20% | 10<br>25% | 11<br>28% |
| Day 2 after surgery | 2<br>5% | 7<br>18% | 8<br>20% | 8<br>20% | 5<br>13% | 10<br>25% |
| Day 3 after surgery | 2<br>5% | 6<br>15% | 8<br>20% | 6<br>15% | 4<br>10% | 14<br>35% |

\* McNemar tests did not find any evidence of a difference from one night or day to any of the next subsequent days ( $p>0.27$ )
† Pain was reported using a visual analog scale.

Overall, up to 87% of patients completed the surveys in the ERAS patient handbooks, though response rates for some questions were as low as 20%. Recalculating percentages by excluding missing data, particularly for discharged patients, showed increased adherence to ERAS recommendations or achievement of desirable outcomes.

## ERAS patient complications

The most common complications included urinary retention or nausea and vomiting (23%, 9/40 patients), and ileus (18%, 7 patients) (**Table 4**). Readmission was rare (8%, 3/40 patients), with one patient readmitted for general weakness, a second for myocardial infarction and a third for pneumonia and surgical related complications. Exactly 60% of patients (24/40) had no complications and only one patient had 3 or 4 complications.

**Table 4:** Complications of 40 ERAS patients*

| Complication | Yes | No |
| --- | --- | --- |
| Nausea or Vomiting | 9<br>23% | 31<br>78% |
| Urinary Retention | 9<br>23% | 31<br>78% |
| Ileus | 7<br>18% | 33<br>83% |
| Wound Dehiscence | 1<br>3% | 39<br>98% |
| Deep Vein Thrombosis | 1<br>3% | 39<br>98% |
| Pneumonia | 1<br>3% | 39<br>98% |
| Wound Infection | 0<br>0% | 40<br>100% |
| Anastomotic Leak | 0<br>0% | 40<br>100% |
| **Number of Patients with...** |  |  |
| No complications | 24 | 60% |
| 1 complication | 7 | 18% |
| 2 complications | 7 | 18% |
| 3 or 4 complications† | 2 | 5% |

\* Three patients (8%) were re-admitted.
† One of these two patients had ileus, nausea or vomiting and urinary retention, while the second patient had these three complications plus wound dehiscence.

### ERAS patient comments

many patients added notes to their handbooks that included comments on the reasons for their responses and about their experience. Retrospective feedback from staff indicated that early removal of catheters was not well-received, particularly in patients that required multiple re-catheterizations. Patients did report that they were highly satisfied with the ERAS procedures, staff and being able to take part in their recovery. Written notes and comments were unanimously positive with respect to patient education and the commitment demonstrated by the RNFA.

### Patient comments

*“Very helpful to have meeting prior to surgery, Useful information on recovery”, “Great to meet JR (RNFA) before surgery – put me at ease”, “Amazed by how quickly I felt good. Day 7 and no pain meds needed”, I feel good going home day 4”, “After my surgery I ate food and passed gas”, “ERAS – amazing”, “I was happy to participate in the program”, “Great care in hospital”, “The program was helpful and informative”*.

### Control vs ERAS Patients

Average age of patients was 70.0 years (standard deviation, SD=12.5) in the control group and 65.4 years (SD=14.0) in the ERAS patient group; this difference was not statistically significant (t-test, p=0.07, mean difference=4.7, 95% confidence interval of the difference −0.3 to 9.6). There was no statistically significant difference in the percentage of females (or males) between the Control group (50.0%) and the ERAS group (45.0%) (Fisher’s exact test, 2-sided p=0.70).

The control group had a significantly higher percentage (69% vs 35%, Fisher’s exact test, p<0.001) of ICD-10-CA code C18 (malignant neoplasm of colon), and significantly lower percentage (0% vs 5%) of C20 (malignant neoplasm of rectum), relative to the ERAS group (**Table 5**). There were no significant differences between control and ERAS groups for 5-character CCI codes (p=0.503) (**Table 6**)(18).

**Table 5:** International Statistical Classification of Diseases and Related Health Problems, 10th Revision, Canada (ICD-10-CA) 3-character code for Control and ERAS patient groups.

| ICD-10-CA * |  | Patient Group † |  | Total |
| --- | --- | --- | --- | --- |
|  |  | Control | ERAS |  |
| C18 | Count | 55 ‡ | 14 § | **69** |
| Malignant neoplasm of colon | % within patient group | 68.8% | 35.0% | **57.5%** |
| C19 | Count | 3 | 0 | **3** |
| Malignant neoplasm of rectosigmoid junction | % within patient group | 3.8% | 0.0% | **2.5%** |
| C20 | Count | 0 § | 2 ‡ | **2** |
| Malignant neoplasm of rectum | % within patient group | 0.0% | 5.0% | **1.7%** |
| D12 | Count | 9 | 8 | **17** |
| Benign neoplasm of colon, rectum, anus and anal canal | % within patient group | 11.3% | 20.0% | **14.2%** |
| K55 | Count | 0 | 1 | **1** |
| Vascular disorders of intestine | % within patient group | 0.0% | 2.5% | **0.8%** |
| K56 | Count | 3 | 1 | **4** |
| Paralytic ileus and intestinal obstruction without hernia | % within patient group | 3.8% | 2.5% | **3.3%** |
| K57 | Count | 4 | 6 | **10** |
| Diverticular disease of intestine | % within patient group | 5.0% | 15.0% | **8.3%** |
| K62 | Count | 1 | 2 | **3** |
| Other diseases of anus and rectum | % within patient group | 1.3% | 5.0% | **2.5%** |
| N32 | Count | 1 | 2 | **3** |
| Other disorders of bladder | % within patient group | 1.3% | 5.0% | **2.5%** |
| Z43 | Count | 4 | 4 | **8** |
| Attention to artificial openings | % within patient group | 5.0% | 10.0% | **6.7%** |
| **Total** | **Count** | **80** | **40** | **120** |
|  | **% within patient group** | **100%** | **100%** | **100%** |

\* The 23 full ICD-10-CA codes were collapsed to ten 3-character codes. Source: CIHI (2015a)
† Fisher's Test (2-sided), Monte Carlo method: $p=0.004$ (95% Confidence Intervals for $p$ : 0.002 to 0.005).
‡ The observed count was significantly higher than predicted by marginal totals.
§ The observed count was significantly lower than predicted by marginal totals.

**Table 6:** Canadian Classification of Health Interventions (CCI) 5-character code for Control and ERAS patient groups.

| CCI code * |  | Patient Group † |  | Total |
| --- | --- | --- | --- | --- |
|  |  | Control | ERAS ‡ |  |
| 1.NM.82 | Count | 4 | 4 | 8 |
| Reattachment, large intestine | % within patient group | 5.0% | 10.8% | 6.8% |
| 1.NM.87 | Count | 75 | 33 | 108 |
| Excision partial, large intestine | % within patient group | 93.8% | 89.2% | 92.3% |
| 1.NQ.74 | Count | 1 | 0 | 1 |
| Fixation, rectum | % within patient group | 1.3% | 0.0% | 0.9% |
| Total | Count | 80 | 37 | 117 |
|  | % within patient group | 100% | 100% | 100% |

\* The full CCI codes were collapsed to three 5-character codes. Source: CIHI (2015b)
† Fisher's Test (2-sided), Monte Carlo method: p=0.503
‡ CCI codes were missing for 3 ERAS patients.

For logarithm-transformed LOS data, Levene’s test of equal variances was significant (F=4.44, p=0.045) and therefore the independent samples Student’s t-test that assumed unequal variances was used. This test found that control patients had a LOS that was significantly longer than ERAS patients (p<0.001) (**Table 7**). The Kolmogorov-Smirnov Test found a significant difference in the distribution of untransformed or transformed LOS between Pre-and Post-ERAS patient groups (p<0.001). The mean difference in logarithm-transformed LOS was 0.548, which was reverse transformed (exponentiated) as a difference of 1.73 days. A test of medians found that median of the control patient group (median LOS=6) was significantly higher than that of the ERAS patient group (median LOS=4) (p=0.001).

**Table 7.** Independent samples t-test of the difference of the mean ln(LOS) between Control and ERAS patient groups.

| Patient Group |  | N | Mean<br>ln(LOS) | Standard Deviation | Standard Error of the<br>Mean |  |
| --- | --- | --- | --- | --- | --- | --- |
| Control |  | 80 | 1.93 | 0.611 | 0.0683 |  |
| ERAS |  | 40 | 1.38 | 0.463 | 0.0732 |  |
| t-test for Equality of Means |  |  |  |  |  |  |
| t | df | p-value (2-tailed) | Mean<br>Difference<br>ln(LOS) | Standard<br>Error of the<br>Difference | 95% Confidence Interval of the Difference |  |
|  |  |  |  |  | Lower | Upper |
| 5.62 | 97.9 | <0.001 | 0.55 | 0.10 | 0.36 | 0.74 |
| Reverse-transformed * |  |  | 1.73 | 1.10 | 1.43 | 2.10 |

\* Values were reverse transformed as $e^x$ , with $e \sim 2.718$ , and $x = \ln(\text{days})$ .

Removing 6 cases in the control group with extreme LOS (≥ 21 days, **Figure 1**) yielded similar statistical test results. A detailed comparison of LOS is provided in **Appendix 1**.

**Figure 1.**
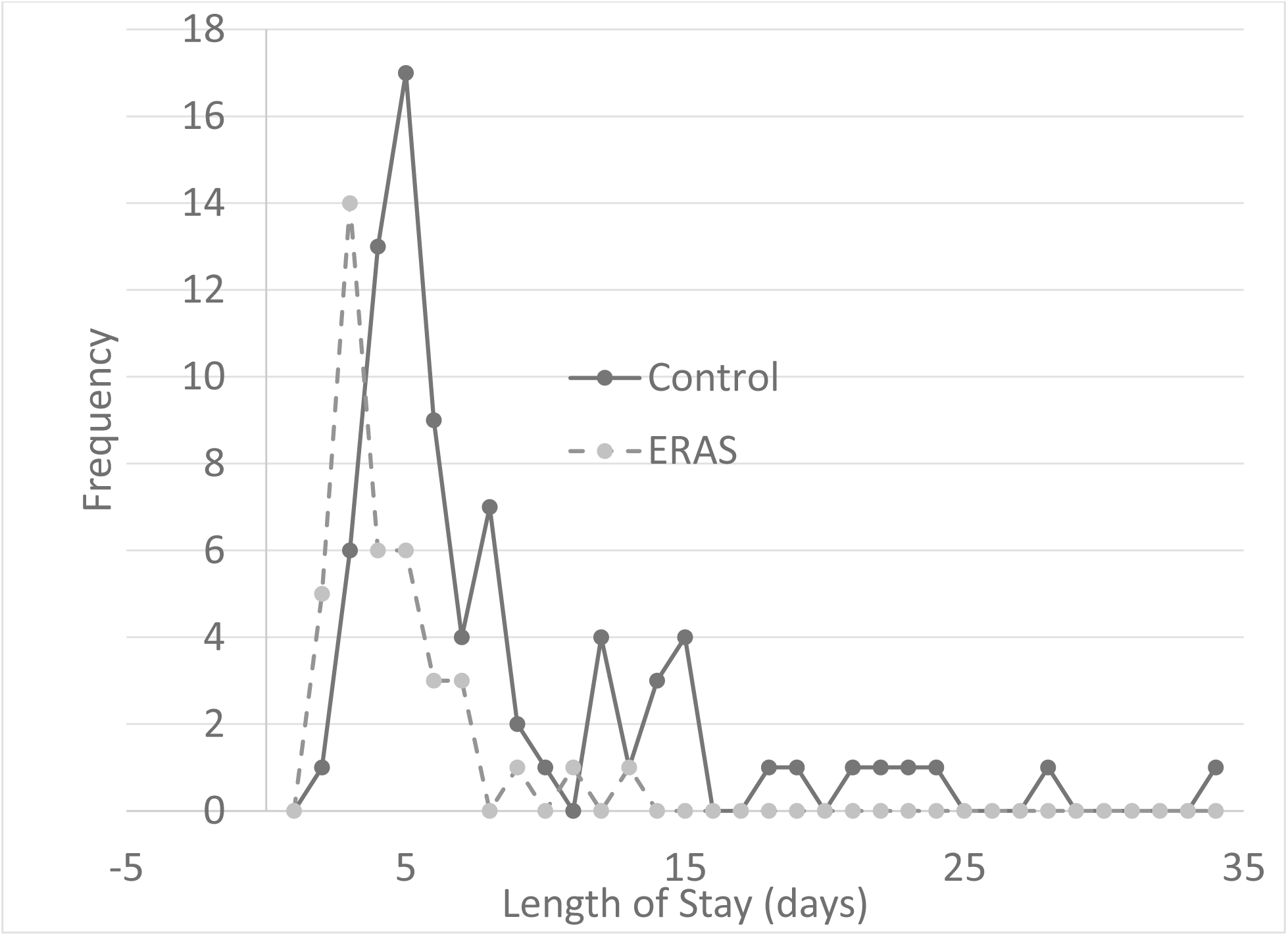
Frequency of length of stay-LOS (days) for 80 Control patients and 40 ERAS patients.

### Results Summary

Most ERAS patients reported adherence to ERAS protocols prior to surgery. Of those who did complete the survey, adherence to the chewing gum protocol, in the days after surgery, was strongest. Most patients were sitting in a chair for their afternoon meal by the first day and were walking down the hallway by the second day. Control patients mean age, male/female percentages and surgical procedures (based on the 3-character CCI code) were not significantly different from that of ERAS patients. Comparison of ICD-10-CA found statistically significant differences between Control group (more malignant neoplasm of colon) and the ERAS group (more malignant neoplasm of rectum). (p≤0.001). The effect of these differences on LOS was not assessed.

All statistical tests on natural logarithm transformed LOS consistently found that Control LOS differed from ERAS LOS. The best available evidence suggests that Control LOS was significantly longer (by 2 days) than ERAS LOS.

## Discussion

ERAS programs using preoperative and postoperative clinical pathways to improve patient outcomes were introduced more than two decades ago however, they are almost exclusively implemented in large urban centers and associated with teaching hospitals. Studies have shown that incorporating ERAS protocols can enhance patient outcomes, reduce the LOS for patients and offer cost savings for the institution (1–7). However, implementing these protocols requires significant multidisciplinary teamwork. Many of the ERAS protocols conflict with traditional practice, which can make uptake difficult. The goal of this project was to demonstrate that ERAS can be performed in a small rural hospital and positively impact patient outcomes.

Over the two-year study, 40 patients were included in the ERAS procedures for colon or rectal surgery. Patients were asked to report on which of the ERAS principles they were informed about and which they complied with. It was found that most patients (75%) consumed the presurgical carbohydrate and chewed gum consistently after surgery. Patient mobility immediately after surgery was also noted, as patients made efforts to both walk in the hallway and take their meals in their chair. Early consumption of food post-operatively was not reported frequently and feedback from patients indicated that the use of an anti-emetic may have improved this. Patients frequently reported feeling nauseous (23%), some to the point of vomiting.

Urinary retention was also high among patients (28%). Comments indicated that early withdrawal of catheters was not well-received, particularly when patients required multiple re-catheterizations.

Patients did, however, report that they were highly satisfied with the ERAS procedures and staff and taking part in their recovery. Feedback in the form of written notes and comments were unanimously positive, particularly with respect to patient education and the commitment demonstrated by the RNFA.

Huntsville, Ontario is a community with an aging population, which was evident in this study. Of the 40 patients treated, the average age was 65 years, equally represented by female and male patients.

Demonstrating improved patient outcomes and a reduced LOS is of importance in this age group as they are predisposed to chronic conditions and susceptible to nosocomial infections.

### Limitations

While all patients were provided the ERAS patient handbook and were asked to complete the handbook throughout their hospital stay, approximately one-quarter did not. Tasking hospital staff, volunteers or research assistants to help patients complete these questionnaires would likely improve response rates, perhaps improve adherence to protocols, and would help identify which ERAS procedures have a higher impact on outcomes.

There are limitations to interpretation based on a matched case study design that uses historical controls. For example, the matching process was conducted on three variables (age, gender and diagnosis) and the effect on LOS of differences between the control and ERAS of these and other variables is unknown. LOS was not adjusted by any method such as the National Surgical Quality Improvement Program risk calculator (ACS 2020). To simplify analyses, the study used unadjusted LOS.

The study was conducted at a single site and results may not necessarily be applicable to other rural hospitals. However, it is worth noting that the ERAS program was successfully implemented in a low-resourced rural hospital, with an aging patient population and compounded by a strong seasonal influx of tourists. Evidence of a reduction in LOS complements success in implementation.

## Conclusions

ERAS consists of a series of preoperative, intraoperative and postoperative clinical pathways aimed at improving clinical care to improve the quality of patient care with patients as active partners in their care. Patient compliance was highest for chewing gum and drinking carbohydrate liquids. Patient outcomes were lowest for ‘peeing on their own’ with several patients requiring re-catheterization. Similarly, the highest complications found in 20% – 30% of patients were urinary retention, nausea and vomiting, and ileus. Pain scores were generally well controlled and overall patient feedback was positive, appreciating that their participation impacted their post-operative recovery. This study found that ERAS could be implemented in a small rural hospital and that LOS could be reduced by two days.

## Future Direction

Creating and implementing pre surgical, surgical and post-surgical electronic order sets for the ERAS pathway is underway at the HDMH. The order sets and training developed through this study are being shared and implemented at the sister site of MAHC, South Muskoka Memoria Hospital.

## Data Availability

All relevant data with statistical analyses are included in the manuscript. Should raw data be requested it is available from the database stored on the Muskoka Algonquin Healthcare server within the ERAS 1 file.

## Abbreviations

CCI: Canadian Classification of Health Interventions
EMR: electronic medical record
ERAS: Enhanced Recovery After Surgery
ICD-10-CA: International Classification of Diseases, 10^th^ revision, Canadian modification
LOS: length of stay
RNFA: Registered Nurse First Assistant
HDMH: Huntsville District Memorial Hospital
MAHC: Muskoka Algonquin Healthcare

## Acknowledgements

This work was supported by the Northern Ontario Academic Medical Association Clinical Innovation Fund. We thank Jennifer Dart and Anca Pengilly for data abstraction, compilation of control cases, assistance in organization and presentation of data for analyses. We also thank the patients and staff of Muskoka Algonquin Healthcare for their participation and support of this study.

## Conflict of interest statement

The authors have no conflict of interest to declare.

## Appendix 1

Length of stay (categories) by sex for control and ERAS patients.

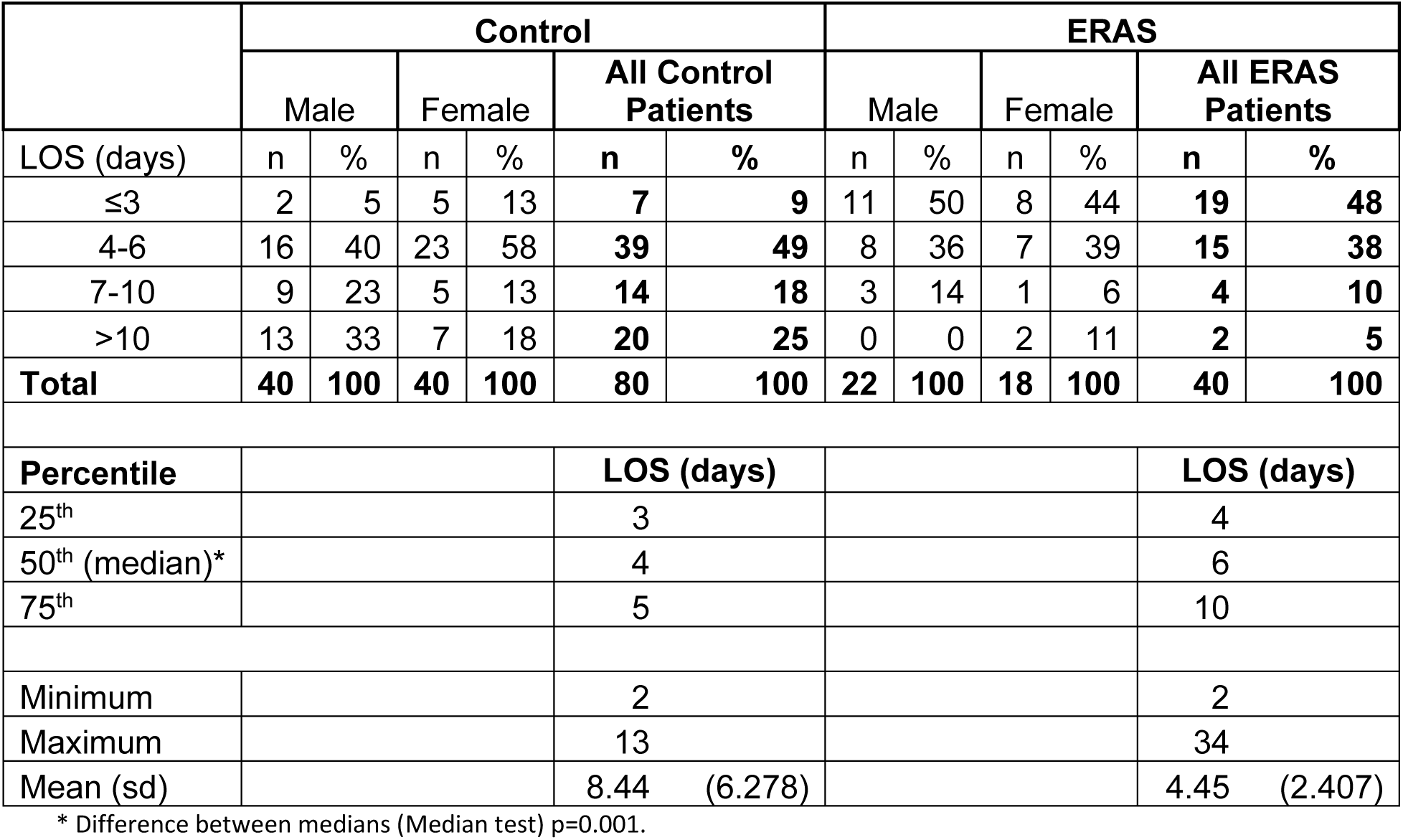

## Notes

**Conflict of Interest:** There are no conflicts of interest to declare.

### Competing Interest Statement

The authors have declared no competing interest.

### Clinical Protocols

N?A

### Funding Statement

This project was funded by the Northern Ontario Academic Medicine Association

### Author Declarations

Laurentian University Research Ethics Board

